# Inherited *STAT3* mutation mediates familial autoimmune disease, leukemogenesis, and responses to immunomodulator

**DOI:** 10.1101/2025.07.08.25331118

**Authors:** Daijing Nie, Qian Zhan

## Abstract

Germline activating mutations in ***STAT****3* result in immune dysregulation, autoimmunity, and increased malignancy risk, yet the precise molecular mechanisms underlying disease heterogeneity remain unclear. We identified a familial germline ***STAT****3* p.R278H mutation shared between the parent diagnosed with T-cell large granular lymphocytic leukemia and the proband presenting with infantile-onset multisystem autoimmune disease-1. Employing whole-exome sequencing, single-cell RNA sequencing, comprehensive cytokine profiling, and flow cytometry-based immunophenotyping, we characterized their distinct clinical phenotypes and underlying immune dysregulation. Single-cell transcriptomics revealed marked expansion of CD8^+^ T cells in the proband, accompanied by altered expression of JAK/STAT pathway genes, including elevated ***STAT3, STAT****1*, and downstream target genes (***BATF, MAF, MYC***), but diminished cytokine transcripts (***IL1B, TNF***). Serum cytokines, however, were markedly elevated, highlighting discordant transcriptional and translational regulation. The parent exhibited additional somatic mutations, notably ***FLT****3*-ITD, suggesting a multi-hit model underpinning leukemogenesis rather than direct oncogenic transformation by ***STAT****3* alone. Immunomodulatory therapies, including cyclosporin A and tacrolimus, controlled autoimmune symptoms in the proband, whereas the parent was refractory to conventional treatments, ultimately succumbing after unsuccessful hematopoietic stem cell transplantation. Our findings emphasize that germline ***STAT****3* activating mutations predispose individuals to complex immune dysregulation and require additional somatic mutations to initiate overt malignancy. This study supports comprehensive genetic profiling and precision medicine approaches for patients harboring congenital ***STAT****3* mutations, aiming to optimize clinical management and improve therapeutic outcomes.

## Introduction

The Janus kinase/signal transducer and activator of transcription (JAK/STAT) signaling pathways play a pivotal role in regulating cytokine-mediated inflammation, immune homeostasis, and oncogenesis. Among the seven identified STAT family members, ***STAT****3* has emerged as a critical mediator, driving potent inflammatory responses and oncogenic processes through persistent activation and aberrant signaling pathways[2, 15, 17, 11]. Numerous studies have underscored the pro-oncogenic role of *STAT3*, which is frequently activated via both intrinsic and extrinsic mechanisms within tumor cells and their immunosuppressive microenvironment [3, 12, 13]. However, direct evidence demonstrating that constitutive intracellular activation of ***STAT****3* alone is sufficient to initiate malignant transformation remains limited. Koskela et al. initially identified somatic ***STAT****3* mutations in 31 of 77 (40%) patients with T-cell large granular lymphocytic leukemia (T-LGLL), all clustered within exon 21 encoding the Src homology 2 (SH2) domain. These mutations resulted in enhanced ***STAT****3* protein activity and transcriptional upregulation of target genes [14]. Subsequent investigations validated these findings and further expanded the mutation spectrum, identifying activating variants outside the SH2 domain, predominantly within the DNA-binding domain [10, 7, 1]. Consequently, ***STAT****3* mutations have become critical diagnostic markers distinguishing T-LGLL from reactive lymphocytosis, given the rarity of such mutations in other malignancies [14]. Nonetheless, experimental evidence exploring the leukemogenic potential of activated ***STAT****3* mutations in T cells remains inconclusive [4, 5]. On the other hand, germline activating mutations of ***STAT****3* represent a distinct entity, characterized as infantile-onset multisystem autoimmune disease-1 (ADMIO1, MIM 615952). First described by Flanagan et al. in 2015, ADMIO1 manifests as an autosomal dominant disorder presenting in early childhood, characterized by autoimmune thrombocytopenic purpura (AITP), autoimmune hemolytic anemia (AIHA), enteropathy, type 1 diabetes mellitus, interstitial lung disease, thyroiditis, short stature, hypogammaglobulinemia, and lymphoproliferation [8, 9, 16]. To date, approximately 28 unique germline ***STAT****3* mutations have been reported in 42 patients, predominantly arising *de novo* and distributed throughout the coding sequence, unlike the restricted hotspots identified in somatic mutations [6].

Despite distinct phenotypic differences between somatic and germline ***STAT****3* mutations, common pathophysiological mechanisms likely exist. To elucidate these mechanisms and phenotype variability, we studied a child aged 5-10 years and one parent carrying an identical heterozygous activating ***STAT****3* mutation but presenting divergent clinical phenotypes. Employing whole-exome sequenc-ing (WES), single-cell RNA sequencing (scRNA-seq), immunophenotyping of lymphocytes, and comprehensive cytokine profiling, we aimed to gain deeper insights into the molecular and cellular underpinnings that govern clinical heterogeneity associated with ***STAT****3* mutations.

### Patient Study and Pedigree Analysis

The proband was a a child aged 5-10 years, born full-term via normal vaginal delivery, without intrauterine or postnatal growth retardation. In early childhood, the proband began to experience recurrent nosebleeds and systemic petechiae. Successive complete blood counts (CBCs) indicated a persistent, marked decrease in platelet count. Initial diagnostic evaluations, including morphologic assessment of bone marrow aspiration smears, led to a diagnosis of immune thrombocytopenic purpura (ITP) at regional hospitals. Systemic corticosteroid treatment initially provided symptom relief; however, glucocorticoid resistance developed within six months. Subsequently, the patient was treated intermittently with gamma globulin infusions and platelet transfusions. One year after the initial diagnosis, ultrasound examinations revealed lymphadenopathy and hepatosplenomegaly. A lymph node biopsy demonstrated reactive follicular hyperplasia. Bone marrow smears exhibited neutropenia, mild megaloblastic changes in granulocytes, and increased cytoplasmic granularity. Throughout the four-year disease course, the patient repeatedly suffered from upper respiratory infections, each episode exacerbating the patient’s thrombocytopenia.

Morphological evaluation of the proband’s bone marrow smears performed in our laboratory revealed normal hyperplasia of granulocytic, erythroid, lymphoid, and megakaryocytic lineages without evident dysplasia or an increased proportion of blasts. However, notable abnormalities included increased vacuolation in monocytes, increased density and size of granules within neutrophils, and occasional neutrophils displaying megaloblastic transformation (Figure 1A, B). Histopathological examination of an enlarged lymph node confirmed reactive follicular hyperplasia (Figure 1C).

**Figure 1:**
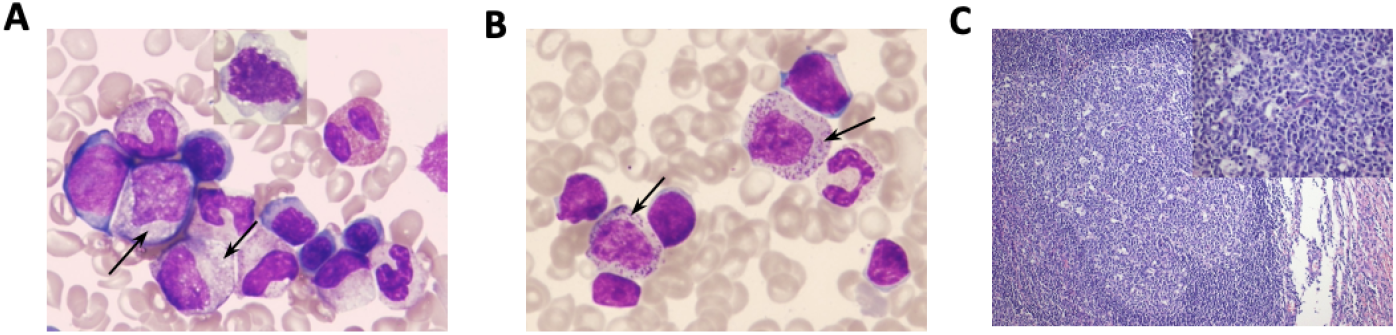
Morphological examination of bone marrow (BM) smear and lymph node (LN) biopsy from the proband. (**A**) BM smear illustrating activated monocytes characterized by increased vacuolation (black arrows). (**B**) BM smear highlighting activated neutrophils with reduced segmentation and prominent granules increased in both density and size (black arrows). Both (**A**) and (**B**) are stained with hematoxylin and eosin (H&E), magnification 100*×*. (**C**) LN biopsy demonstrating reactive follicular hyperplasia (low magnification) and follicle center cell morphology (high magnification). H&E staining, original magnifications 10*×* (left) and 40*×* (right).

One of the proband’s parents, initially presenting in young adulthood with fatigue, demonstrated severe anemia and lymphocytosis without neutropenia or thrombocytopenia in CBC analyses. Further clinical evaluations identified moderate splenomegaly and mild hypergammaglobulinemia. Bone marrow aspiration smears and biopsy specimens indicated marked erythroid hypoplasia, with normal cellularity in other hematopoietic lineages. Flow cytometric immunophenotyping consistently identified a predominant population of T cells characterized by CD5^*−*^/low, CD2^+^, CD3^+^, CD8^+^, CD11c^+^, and CD57^+^ expression, along with T-cell receptor (TCR) v positivity in repeated assays from 2014 to 2016. Clonality of the T-cell population was confirmed by polymerase chain reaction (PCR)-based assays demonstrating detectable TCR v rearrangements. Consequently, the parent was diagnosed with T-cell large granular lymphocytic leukemia (T-LGLL) accompanied by pure red cell aplasia (PRCA).

To elucidate the disease-causing mutations in the proband, we performed WES and identified a heterozygous ***STAT****3* mutation (c.833G*>*A, p.R278H).

This mutation was classified as pathogenic according to the American College of Medical Genetics and Genomics (ACMG) guidelines. The clinical manifestations associated with germline ***STAT****3* activating mutations recorded in the Online Mendelian Inheritance in Man (OMIM) database closely resemble those observed in the proband, including early-onset autoimmune diseases, lymphadenopathy, and hepatosplenomegaly.

Further analysis of the parent’s WES data revealed the identical heterozygous ***STAT****3* R278H mutation along with an additional heterozygous ***TAL****1* mutation (c.184G*>*C, p.G62R; rs743269), whose clinical significance remains uncertain. The ***TAL****1* gene encodes a transcription factor expressed in normal hematopoietic stem cells, progenitor cells, erythrocytic, and megakaryocytic lineages. ***TAL****1* is typically transcriptionally silenced during lymphocyte development; however, aberrant expression can arrest T-cell differentiation at the late cortical stage and facilitate oncogenesis. The oncogenic mechanism attributed to ***TAL****1* usually involves abnormal expression resulting from chromosomal translocations, intrachromosomal rearrangements, or enhancer mutations. It remains unclear whether missense mutations alone, such as the observed ***TAL****1* G62R mutation, can induce ectopic expression or trigger oncogenesis.

Tumor initiation typically arises from the accumulation of multiple genetic alterations, granting cells proliferative and survival advantages. Hence, somatic mutations are essential in this context. The parent developed leukemia in late young adulthood, prompting us to question the potential roles of germline ***STAT****3* and ***TAL****1* mutations in neoplastic transformation. Deep panel sequencing and capillary electrophoresis identified a somatic FLT3 internal tandem duplication (FLT3-ITD), involving a 24-bp insertion with a variant allele frequency (VAF) of 28%. The ***FLT****3* gene encodes a receptor tyrosine kinase critical for hematopoiesis; mutations in this gene are observed in approximately 1–3% of acute lymphoblastic leukemia (ALL) patients.

In this pedigree, we confirmed the ***TAL****1* G62R mutation was inherited, whereas the ***STAT****3* R278H mutation occurred de novo in the parent by Sanger sequencing. Familial biological relationships were verified by short tandem repeat (STR) analysis using the AmpFLSTR Identifiler kit (Thermo Fisher, CA, USA), examining 15 STR loci. (Data are available upon request from the corresponding author.)

### *STAT3* mutation leads to Immune Dysregulation

In order to investigate the effect of the ***STAT****3*, we performed scRNA-seq on the peripheral blood mononuclear cells of the proband. A total of 9,114 individual cells were sequenced, generating an average of approximately 6,000 transcripts derived from around 1,200 genes per cell. After removing outliers, we further analyzed 8,660 cells. Using t-distributed stochastic neighbor embedding (t-SNE), we identified nine distinct cell clusters (Fig. 2A). Each cluster was annotated based on the expression of specific marker genes (Figure 2B). The most abundant cell type in the proband’s peripheral blood mononuclear cells (PBMCs) was CD8^+^ T cells, including both effector and naïve subsets, accounting for 44.73%. This was followed by CD14^+^ monocytes (18.2%), CD4^+^ T cells (14.98%), CD19^+^/CD20^+^ B cells (14.16%), and natural killer (NK) cells (5.66%) (Fig. 2C). Attempts to further delineate subsets within CD4^+^ T cells, such as Th1, Th2, Th17, and Treg cells, were unsuccessful due to their scarcity and extremely low expression levels of characteristic markers.

**Figure 2:**
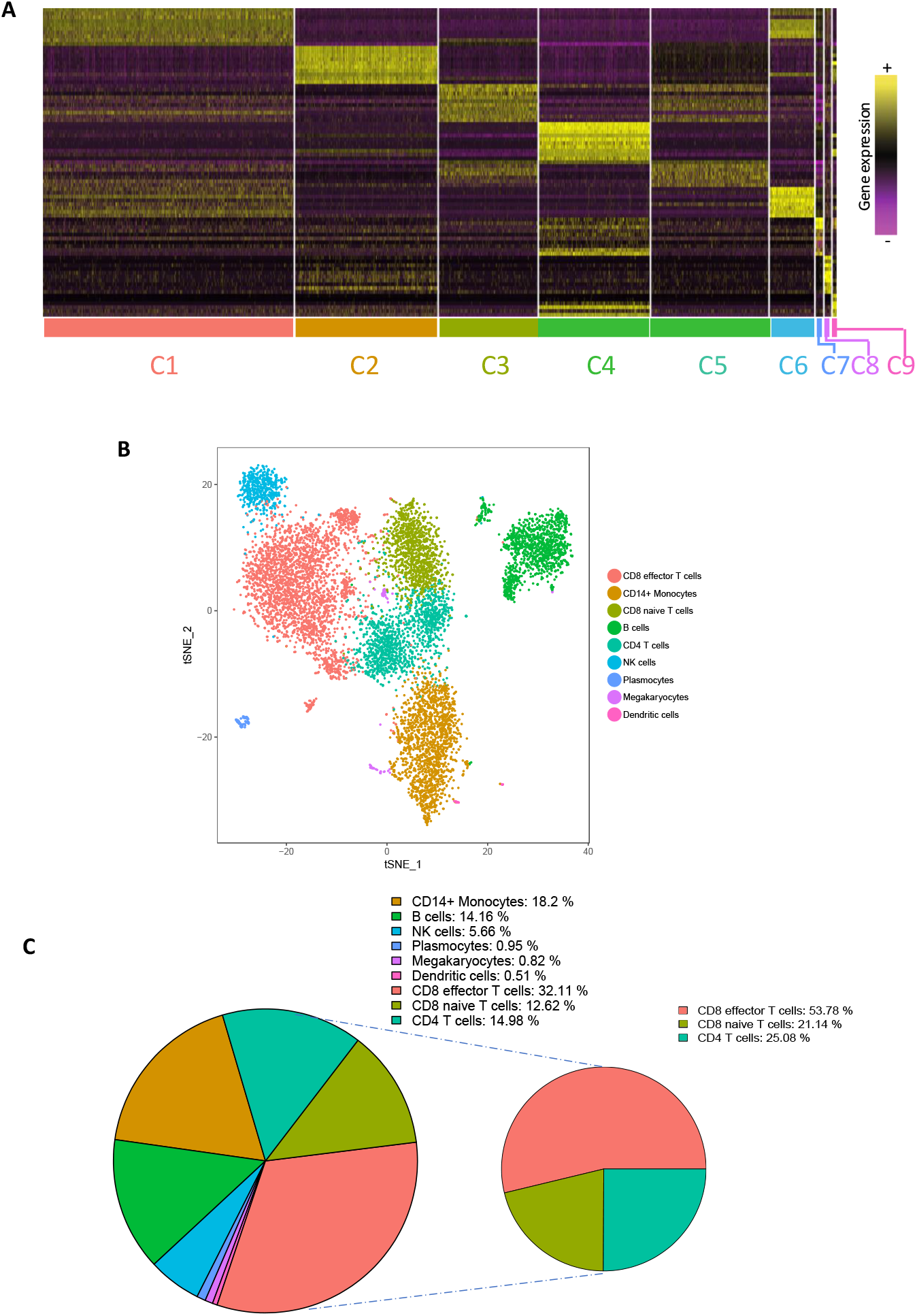
Single-cell RNA sequencing (scRNA-seq) reveals distinct cell clusters and distribution in the proband’s peripheral blood mononuclear cells (PBMCs). Heatmap demonstrating expression profiles across nine distinct cell clusters, with assigned cluster identities annotated at the bottom. (**B**) t-distributed stochastic neighbor embedding (t-SNE) visualization illustrating distinct PBMC clusters, color-coded accordingly. Cluster identities are labeled adjacent to each cluster. (**C**) Bar plot showing the proportion and distribution of cells across each identified cluster.

We next performed comparative gene expression analysis between the proband and the genderand agematched healthy control, focusing particularly on target genes within the JAK/STAT3 pathway, including cytokines, transcription factors, and related regulatory genes such as ***SOCS****3*, ***TIM****P1*, and ***MYC***. The expression of ***SOCS****3* was not significantly different between the proband and control (Figure 3). However, other STAT3 target genes, including ***BATF, MAF***, and ***MYC***, demonstrated significantly elevated expression levels across most clusters in the proband. Additionally, cytokine profiling within CD4^+^ T cells revealed significantly reduced expression of ***IL1B*** and ***TNF*** (both with *p* = 0), as well as decreased, though less dramatically, expression of ***IL6*** and ***IFNG***. Among genes related to the JAK/STAT signaling pathway, the expression of ***JAK****3*, ***STAT****1*, and ***STAT****3* was significantly elevated in the proband across five analyzed clusters.

**Figure 3:**
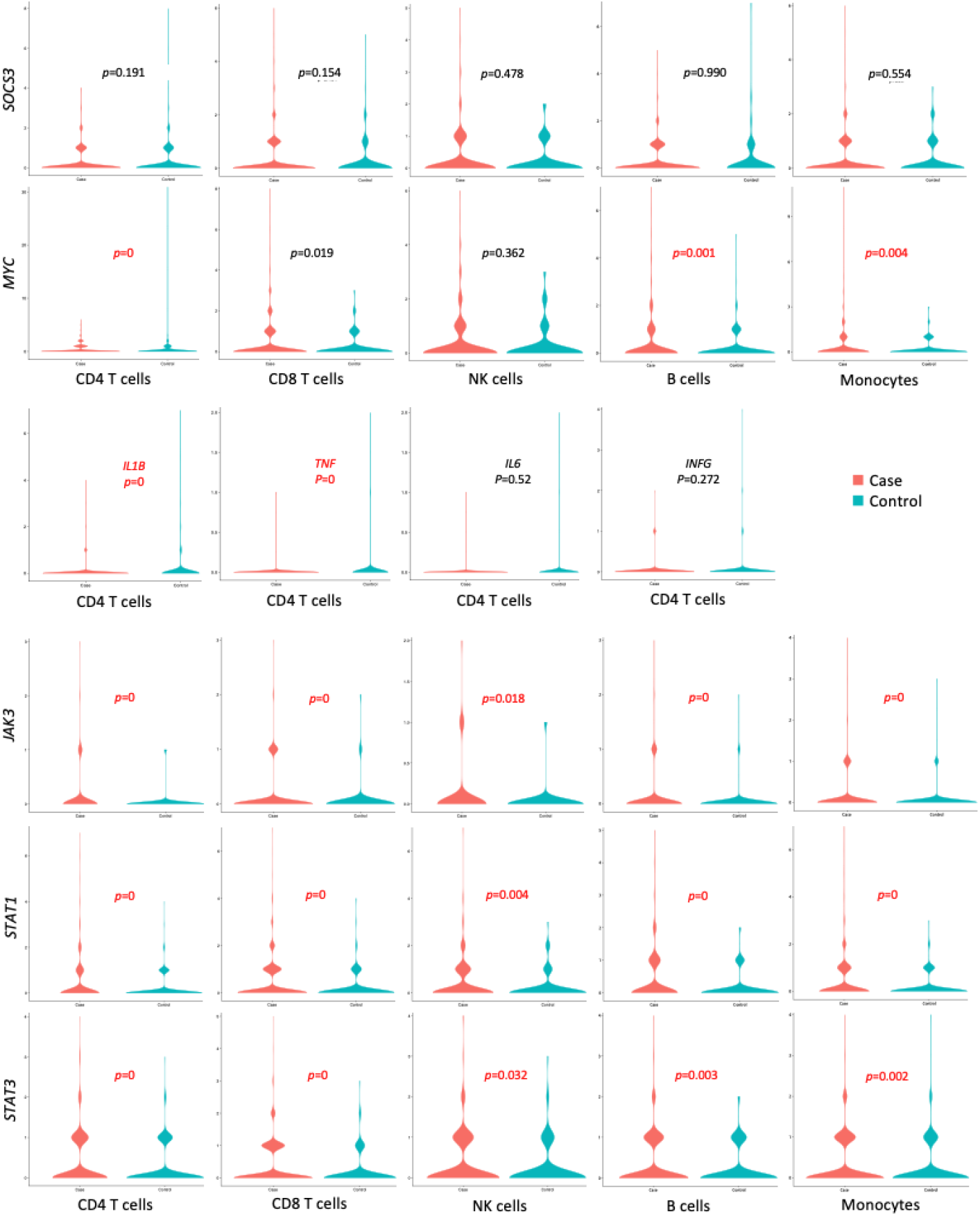
Violin plots comparing gene expression profiles between the proband (red) and healthy control (green). Genes analyzed include *STAT3* target genes, cytokines, and members of the JAK/STAT signaling pathways within immuno-cytes. Significantly elevated gene expressions in the proband are indicated by highlighted red *p*-values.

Previous studies indicated a heterogeneous immunophenotype in patients harboring germline activating mutations of ***STAT****3*. In our proband, the overall lymphocyte count was elevated, comprising 60.5% of peripheral blood mononuclear cells (PBMCs; reference range: 19.85–40.56%). The increase was most pronounced among CD3^+^CD8^+^ T cells, whereas CD16^+^/CD56^+^ NK cell counts were reduced (Figure 4A–C). These findings were consistent with the single-cell RNA-seq results. In contrast to previous observations, no significant increase was noted in CD3^+^CD4^−^CD8^−^ or CD3^+^CD4^+^CD8^+^ T-cell populations (Figure 4D), suggesting intact mechanisms of positive and negative thymic selection. Within the CD3^+^CD4^+^ T-cell compartment, CD183^low/−^CD196^−^ Th2 cells pre-dominated, whereas counts of CD183^−^CD196^+^ Th17 cells and CD4^+^CD25^+^CD127^dim^ regulatory T cells (Treg) remained within normal ranges (Figure 4E–F).

**Figure 4:**
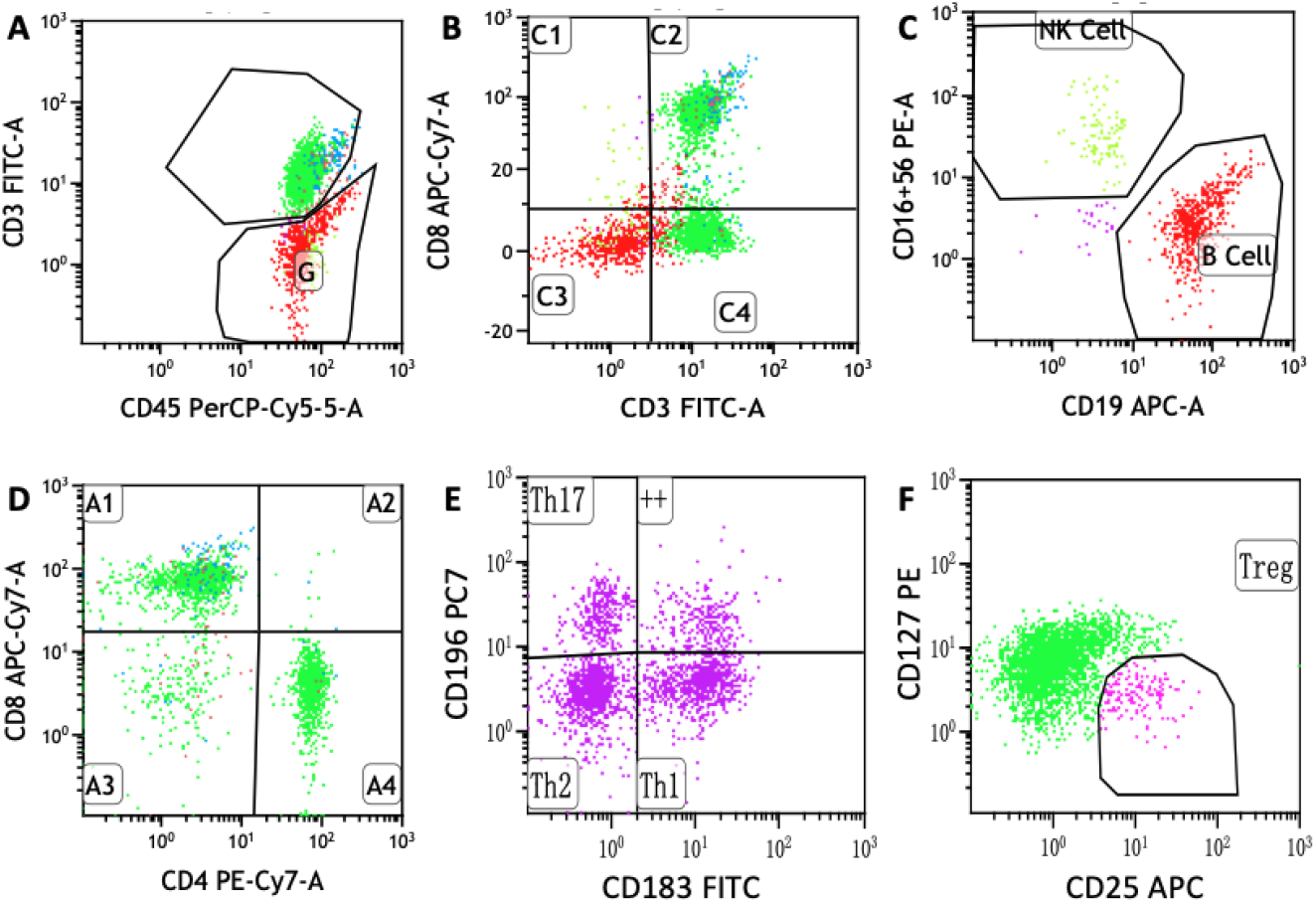
Flow cytometric immunophenotyping of lymphocyte subsets in the proband. (**A**) CD3 and CD45 staining demonstrates that CD3^+^ T cells constitute 80.26% of gated lymphocytes (reference range: 24.93–81.56%). (**B**) CD3^+^CD8^+^ T cells comprise 48.63% of gated lymphocytes (reference range: 16.40–66.76%). (**C**) CD16^+^/CD56^+^ NK cells represent 2.71% (reference range: 8.3–25.6%), and CD16^+^ B cells account for 15.57% (reference range: 5.0–39.5%) of gated lymphocytes. (**D**) CD3^+^CD4^−^CD8^−^ double-negative T cells represent 6.75% (reference range: 0–12.0%), and CD3^+^CD4^+^CD8^+^ double-positive T cells represent 0.63% (reference range: 0–2.0%) of gated lymphocytes. (**E**) CD196 and CD183 staining within CD4^+^ T-cell subsets reveals the following proportions: CD183^+^CD196^low/−^ Th1 cells (34.54%), CD183^low/−^CD196^−^ Th2 cells (40.06%), and CD183^−^CD196^+^ Th17 cells (13.31%). (**F**) Regulatory T cells (CD4^+^CD25^+^CD127^dim^) constitute 4.45% of CD4^+^ T cells (reference range: 2.01–8.2%).

We also assessed serum concentrations of twelve cytokines. Interestingly, despite the decreased intracellular cytokine transcription observed, most serum cytokines were moderately elevated. Notably, IL-1*β*, IL-5, TNF-*α*, and IFN-*γ* exhibited the most prominent elevations, increasing 8.3-, 7-, 4.23-, and 3.7-fold, respectively, above reference ranges (data not shown).

### Clinical Management

Following the confirmed diagnosis of ADMIO1, immunosuppressive therapy was initiated for the proband. Cyclosporin A (CsA) effectively maintained platelet counts between 166 *×* 10^9^/L and 302 *×* 10^9^/L; however, lymphadenopathy and hepatosplenomegaly persisted without improvement. Eleven months into CsA therapy, the dose was reduced by one-third due to severe gingival hyperplasia, which resulted in recurrence of purpura and a rapid drop in platelet count within ten days. Subsequently, tacrolimus was administered, leading to stable normalization of platelet counts over the following three months and resolution of lymphadenopathy, hepatomegaly, and splenomegaly at the most recent followup.

In contrast, over the three-and-a-half-year course of illness, the parent exhibited resistance to both CsA and methotrexate (MTX). Furthermore, one course of fludarabine induced severe bone marrow suppression. Due to persistently el-evated levels of circulating HLA-I and HLA-II antibodies, platelet transfusions were effective only following plasmapheresis. Unfortunately, the parent eventually succumbed to pulmonary and gastrointestinal infections following failed maternal HLA-haploidentical hematopoietic stem cell transplantation (HSCT).

## Discussion

In this study, we identified a familial activating mutation, ***STAT****3* R278H, shared by a parent diagnosed with T-cell large granular lymphocytic leukemia (T-LGLL) and his child, who presented with ADMIO1 characterized by autoimmune manifestations. Previous reports have described the ***STAT****3* R278H mutation in association with early-onset autoimmune disorders, hemolytic anemia, lymphoproliferation, and immunodeficiency, linking this mutation directly to increased phosphorylation and enhanced transcriptional activity of STAT3 target genes [8, 16]. Consistently, we demonstrated widespread and persistent inflammatory activation involving granulocytes, monocytes, and lymphocytes, alongside the upregulation of ***STAT****3* and its downstream target genes, providing further molecular validation of hyperactivated JAK/STAT signaling in our proband.

Despite harboring the same germline mutation, significant clinical heterogeneity between the parent and child underscores the complexity of phenotypic expression associated with activating ***STAT****3* variants. Single-cell RNA-seq and immunophenotyping data indicated a marked elevation in CD8^+^ T-cell populations within the proband, accompanied by divergent cytokine expression profiles at transcriptional and protein secretion levels. Notably, elevated serum cytokine levels, including IL-1*β*, TNF-*α*, IL-5, and IFN-*γ*, contrasted sharply with reduced intracellular cytokine transcripts, suggesting differential post-transcriptional regulatory mechanisms or altered cytokine secretion dynamics in the context of constitutively activated STAT3. Such discrepancies highlight the necessity of careful interpretation of cytokine profiling in autoimmune and inflammatory disorders.

STAT3 signaling is intricately balanced with other members of the JAK/STAT pathway, particularly STAT1, which exhibits compensatory upregulation in response to aberrant STAT3 activation [18, 16]. This compensatory mechanism was clearly observed in our study through the significantly increased STAT1 expression. Interestingly, although previous literature suggested that activating STAT3 mutations might decrease STAT1 phosphorylation levels, our data indicate that transcriptional upregulation of STAT1 could represent an adaptive cellular attempt to restore signaling equilibrium, potentially impacting disease progression and clinical outcomes.

In terms of oncogenesis, germline activating ***STAT****3* mutations, although strongly associated with autoimmune and lymphoproliferative phenotypes, rarely induce malignant transformation in isolation. The present study provides additional evidence that tumorigenesis in patients harboring constitutional STAT3 activation likely requires secondary genetic aberrations or environmental influ-ences. Indeed, the parent, who developed T-LGLL, possessed additional genetic alterations, including a germline ***TAL****1* G62R variant and an acquired somatic FLT3-ITD mutation, each potentially conferring proliferative and survival advantages to leukemic cells. The oncogenic role of TAL1 typically results from aberrant expression due to structural genomic abnormalities rather than isolated missense mutations; thus, the contribution of the ***TAL****1* G62R variant to leukemogenesis remains uncertain but warrants consideration in combination with other genetic lesions. Notably, FLT3-ITD mutations are well-established drivers in acute leukemias, enhancing cellular proliferation and survival through constitutive receptor tyrosine kinase signaling. Collectively, these genetic alterations may reflect the cumulative burden necessary to transition from chronic lymphoproliferation and immune dysregulation to overt malignancy.

Familial occurrences of T-LGLL are exceedingly rare, with few documented cases involving somatic STAT3 mutations restricted mainly to the SH2 domain (exon 21). Our identification of a familial germline ***STAT****3* mutation outside this canonical region (exon 9, encoding the coiled-coil domain) expands the known spectrum of pathogenic STAT3 variants, suggesting that comprehensive genetic screening beyond traditional mutation hotspots is essential to capture clinically relevant alterations[10]. Moreover, the considerable phenotypic variability observed, even among individuals harboring identical germline STAT3 mutations, emphasizes the impact of genetic modifiers, environmental triggers, and potentially other yet unidentified somatic mutations.

Therapeutically, ADMIO1 poses significant clinical management challenges. Traditional immunosuppressive therapies, including corticosteroids, cyclosporin A (CsA), and tacrolimus, often yield partial and transient responses, necessitating a transition to more targeted therapies. Encouraging outcomes from IL6 receptor blockade (tocilizumab) have recently emerged, suggesting potential utility as targeted therapy in ADMIO1 management. However, caution is advised regarding HSCT, given the high rate of adverse outcomes and transplant-related morbidity and mortality reported to date [6]. The parent’s refractory clinical course, resistance to multiple immunosuppressive agents, chemotherapy intolerance, and eventual HSCT failure further underscore the complexity and therapeutic dilemma inherent to managing T-LGLL with underlying STAT3 dysregulation. It highlights the necessity for precision medicine approaches, guided by comprehensive genetic and immunophenotypic profiling, to improve therapeutic efficacy and minimize adverse outcomes.

Our study is limited by the unavailability of fresh samples from the parent, precluding a matched analysis of single-cell transcriptomics and comprehensive immunophenotyping to further delineate cellular and molecular differences underpinning distinct disease trajectories. Nevertheless, the detailed genetic characterization performed provides valuable insights into the natural history and progression from congenital immune dysregulation to hematologic malignancy. Future longitudinal studies incorporating multi-omics approaches, coupled with rigorous functional validation, are critical for elucidating the complex interplay between germline activating STAT3 mutations, immune dysregulation, and leukemogenesis.

In conclusion, our integrative analysis delineates the heterogeneous clinical outcomes associated with germline activating ***STAT****3* mutations, underscoring their role as necessary but not sufficient oncogenic drivers. The identification of additional genetic aberrations highlights a multi-hit model of leukemogenesis, emphasizing the importance of comprehensive genetic profiling for informed clinical management. Our findings advocate for a precision medicine approach, leveraging targeted therapies and meticulous risk stratification to improve clinical outcomes in patients with inherited ***STAT****3* mutations.

## Data Availability

All data produced in the present study are available upon reasonable request to the authors

## Declaration

Written informed consent was obtained from the statutory guardian of the proband and all participating family members in accordance with the Declaration of Helsinki. Ethical approval for the study was granted by the Ethics Committee of Hebei Yanda Lu Daopei Hospital. The authors Declare that there is no conflict of interest in this study.

## Notes

### Competing Interest Statement

The authors have declared no competing interest.

### Funding Statement

This study did not receive any funding

### Author Declarations

The Ethics Committee of Hebei Yanda Lu Daopei Hospital gave ethical approval for this work.

